# Study protocol for a randomized clinical pilot trial investigating feasibility and efficacy of augmenting a virtual reality-assisted intervention targeting auditory verbal hallucinations with biofeedback: the Neuro-VR study

**DOI:** 10.1101/2025.09.19.25336131

**Authors:** Amalie Fabricius Habla, Sara Breivik Soleim, Anne Sofie Due, Tina Højsgaard Tinglef, Kasper Eskelund, Júlia Díaz-i-Calvete, Kit Melissa Larsen, Tina Dam Kristensen, Bjørn H. Ebdrup, Merete Nordentoft, Daniel Lyngholm, Kamilla Woznica Miskowiak, Karen Sandø Ambrosen, Louise Birkedal Glenthøj

## Abstract

**Introduction:** Auditory Verbal Hallucinations (AVH) are among the most frequent and severe symptoms in schizophrenia and related psychotic disorders. Virtual Reality (VR)-assisted interventions have emerged, demonstrating promising potential in reducing AVH severity. This treatment approach may be challenged with regards to feasibility, particularly when therapeutically managing the anxiety-related reactions associated with AVH. This pilot study evaluates the feasibility and acceptability of augmenting VR-assisted therapy with real-time biofeedback to address these challenges. The integration of biofeedback enables continuous adaptation of therapy based on physiological responses while allowing participants to train self-regulation of these parameters.

**Methods:** Neuro-VR is a randomized clinical pilot trial utilizing a mixed-methods design. Thirty participants with schizophrenia spectrum disorders and AVH will be randomized to either eight sessions of VR-assisted therapy or eight sessions of VR-assisted therapy augmented with real-time biofeedback. Assessments will be conducted at baseline and post-treatment. Outcome measures include both clinical metrics, electroencephalogram recordings, and qualitative interviews to evaluate feasibility, acceptability, and potential treatment effects of the combined approach.

**Discussion:** This study will explore whether integrating biofeedback into VR-assisted therapy enhances personalization, supports emotion regulation, and improves tolerability. The findings will provide preliminary evidence on the utility of physiological markers to guide VR-based interventions for AVH and inform the development of individualized, effective treatments for patients with schizophrenia.

**Trial registration:** ClinicalTrials.gov, NCT06628323 (https://clinicaltrials.gov/study/NCT06628323). Registered August 19, 2024.

## Introduction

### Auditory Verbal Hallucinations in schizophrenia

Schizophrenia and related psychotic disorders are severe mental illnesses that are characterized by psychotic and negative symptoms, as well as cognitive deficits [1]. Auditory Verbal Hallucinations (AVH), referring to the perception of hearing voices without external stimuli, are among the most prevalent symptoms of schizophrenia, affecting approximately 75% of the clinical population [2,3]. Although the content of AVH is heterogeneous and influenced by cultural factors, individuals diagnosed with schizophrenia and AVH often report voices with negative content, such as criticizing or threatening voices [4–7]. This type of AVH often causes severe distress to the individual and disruption to daily and social functioning [5,8,9]. Psychotic symptoms remain resistant to antipsychotic treatment in approximately 30% of the affected individuals [10,11], underscoring the need for alternative treatment options, such as psychotherapeutic interventions, specifically targeting AVH [12].

### Virtual Reality-assisted psychotherapy for AVH

Cognitive Behavioural Therapy (CBT) is considered as the gold standard psychotherapeutic intervention for psychosis. However, meta-analyses reveal only small to moderate effect sizes of this approach [12–14]. This shortcoming has motivated research aimed at developing targeted and potentially briefer therapies for AVHs [15]. Several novel approaches conceptualize hallucinated voices as identity-bearing entities with whom the individuals hearing voices, maintain a personal relationship [12,16,17]. A major aspiration of these more efficient approaches is to address the power dynamics within this relationship, aiming to enhance the individual’s feeling of control towards the voice [12,17,18]. The so-called AVATAR therapy is a part of this field of relational approaches, which utilizes technology to create a digital representation of the distressing voice, termed an avatar [15,19]. The avatar is created to visually correspond to the participant’s experience of the perceived hallucinated voice, and to auditorily match the voice heard by the participant by transforming the voice of the therapist using a computer program. The therapist facilitates dialogues between the participant and avatar, enabling a tangible unfolding of the power dynamics between the two parts in a controlled environment. Early studies on AVATAR therapy such as the pilot study by Leff et al. [19], as well as the following AVATAR 1 and 2 trials [15,20], utilized a computerized system to deliver the therapy. This intervention has been further developed in subsequent studies involving immersive Virtual Reality (VR)-technology [21,22]. Findings from these studies suggest that this therapeutic approach has promising potential in reducing the severity of AVH. Building on this work, we have previously evaluated the efficacy of a fully immersive VR-assisted intervention targeting AVH in the Challenge-trial, a large-scale randomized clinical trial [23], finding it to be an effective and feasible treatment approach [24,25].

### Potential challenges in VR-assisted therapy for AVH

The visual personification of the hallucinated voice, involving the delivery of its verbatim content through the voice of the avatar, offers the potential to create a realistic experience of engaging in a confrontation with the voice [26]. In the context of VR-assisted therapy, the term *presence* is commonly used to capture this individual experience of being present or “being there” in the digital environment [27,28]. Rus-Calafell et al. [26] investigated the impact of the participants’ sense of voice presence on the outcomes of AVATAR therapy, finding that mid to high levels of presence were reported consistently through the therapy sessions. This indicates that AVATAR therapy is effective in its aim of delivering a realistic experience of being in a dialogue with the hallucinated voice. Furthermore, they found that the *interaction* between presence and a reduction in the participant’s anxiety throughout therapy sessions was associated with improvements of voice severity and frequency post treatment. This suggests that the therapy outcome may be influenced by the combination of participants feeling less anxious over time, in the context of a realistic simulation of the voice. In light of this, the activation of fear- and anxiety responses related to the auditory hallucination may be considered an important aspect of the therapy, by potentially providing the participant with the opportunity to manage the voice-related emotional distress in new ways [18,26]. However, this activation may involve challenges during therapy. Ward et al. [18] present an account of therapeutic targets in AVATAR therapy based on a case review from completed interventions in the AVATAR trial. Notably, all therapy withdrawals occurred during the initial phase, involving direct exposure to the verbatim content from the avatar [18]. This highlights the potential difficulty of this phase for both therapists and the participants. Considering these experiences, determining the optimal level of exposure emerges as a both critical and challenging aspect of delivering VR-assisted therapy for AVH.

### Biofeedback

Biofeedback is a technique that uses biosensors and electronic devices to monitor and display physiological responses in real time, helping the patient modify their bodily reactions [29,30]. These physiological signals provides objective measures, thereby enabling continuous monitoring of participants’ physiological responses [31]. Among the most relevant signals are those reflecting autonomic nervous system activity, which comprises both sympathetic and parasympathetic branches. Sympathetic activation occurs rapidly in response to perceived threats, triggering the classic “fight or flight” response. In contrast, the parasympathetic nervous system plays a key role in restoring the body to a calm, relaxed state following arousal [32]. Heart rate variability (HRV) and heart rate (HR) are prominent biomarker within biofeedback research [33,34]; HR being the number of heart beats per minute and HRV, the fluctuation of the length of heart beat intervals [35]. Reduced HRV reflects diminished parasympathetic activity and is consistently associated with elevated stress and anxiety levels [35,36]. Moreover, increases in heart rate (HR) show strong correlations with anxiety [37], making HR a suitable parameter for real-time monitoring [38].

### Combining VR and biofeedback

By combining VR and biofeedback, the study benefits from VR’s capacity to immerse participants in a realistic environment, while simultaneously providing both the participant and therapist with real-time physiological data relevant for treatment progress. Previous studies have shown successful implementation of VR and biofeedback in treating conditions such as anxiety [29], stress [39] and pain [40]. Although an ongoing study explores neurofeedback combined with VR in schizophrenia, its design and therapeutic approach differ from ours [41]. To the best of our knowledge, this is the first study to investigate a biofeedback-augmented VR-assisted intervention specifically targeting AVH.

### Objectives

The primary objective of the study is to examine the feasibility and acceptability of augmenting a VR-assisted intervention with real-time biofeedback for treating AVH in schizophrenia. Secondary objectives are to explore preliminary evidence of enhanced therapeutic efficacy resulting from biofeedback integration and to investigate potential neurophysiological mechanisms underlying treatment outcomes through electroencephalogram (EEG) assessments conducted at baseline and post-treatment.

We hypothesize that, in patients with schizophrenia:

1. VR-assisted therapy augmented with biofeedback will be feasible and acceptable.
2. VR-assisted therapy augmented with biofeedback will provide preliminary evidence of superiority over VR-assisted therapy alone, in reducing the severity of AVH and improving daily functioning.

## Materials and methods

### Study design

The study is carried out at the Mental Health Centre Copenhagen, Mental Health Services, Capital Region of Denmark. It is a two-arm pilot randomized clinical trial (RCT) that utilizes a mixed-methods design combining quantitative and qualitative methods. A total of 30 participants diagnosed with a schizophrenia spectrum disorder based on ICD-10 (codes: F20, 22-23, 25-29) from the psychiatric outpatient facilities in the Capital Region of Denmark and the Region Zealand will be enrolled. The participants will be randomly assigned to one of the two treatment arms:

1. Control group: 8 sessions of VR-assisted therapy
2. Experimental group: 8 sessions of VR-assisted therapy augmented with real-time biofeedback

Participants will be assessed at baseline and at treatment cessation (12 weeks post baseline) (Fig 1). In addition to clinical evaluations, both time points will include EEG assessments to examine the intervention’s potential effects on brain activity.

**Fig 1.**
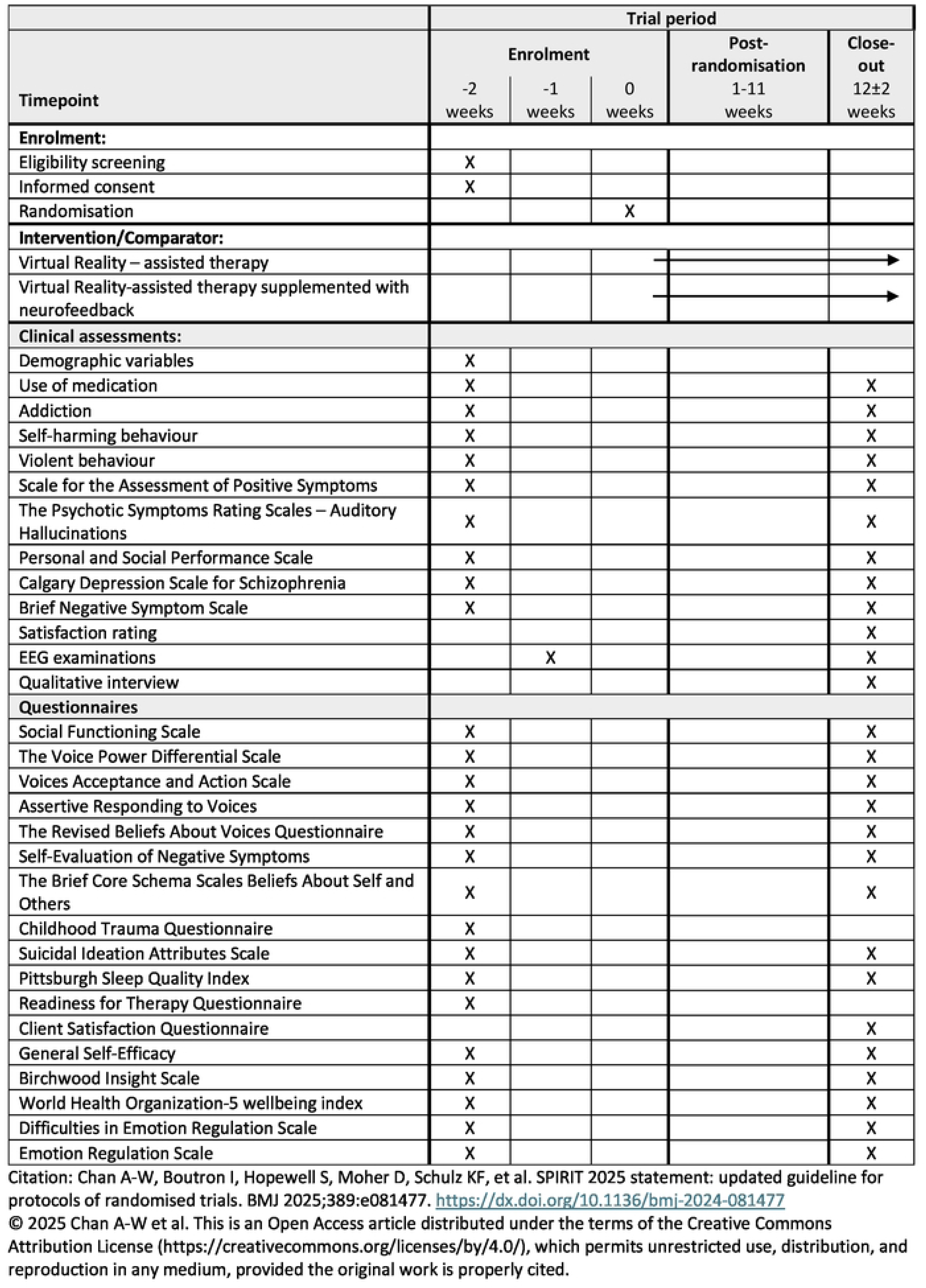
SPIRIT schedule of enrolment.

A mixed-methods design is employed to provide a comprehensive understanding of feasibility and acceptability. Qualitative methods are used to capture the complexity of factors influencing these outcomes [42,43], incorporating perspectives from both patients and therapists. These insights will help refine the intervention and inform potential future clinical implementation.

The inclusion period started on 12 November 2024 and is ongoing. The final participant is expected to be included by 1 November 2025, with data collection anticipated to be completed by 1 February 2026.

Results are expected by June 2026.

### Participants

Participants must meet the following eligibility criteria:

Inclusion criteria:

- Willing and able to give informed consent
- Age 18-65
- Diagnosed with a schizophrenia spectrum disorder (ICD-10: F20, 22-23, 25-29)
- Auditory verbal hallucinations within the past 3 months (corresponding to a SAPS score on auditory hallucinations of ≥ 3)
- No changes in antipsychotic medication within the past 4 weeks and no planned changes in antipsychotic medication in the 12 weeks following inclusion in the study

Exclusion criteria:

- Diagnosis of organic brain disease
- Current diagnosis of substance dependance hindering engagement in therapy
- Intellectual disability (IQ < 70)
- A command of Danish or English insufficient to undergo therapy
- Hearing voices in a language that the therapist does not speak
- Unable to identify a dominant voice to work with

### Sample size

The primary objective of this pilot study is to assess the feasibility and acceptability of the experimental intervention and study protocol, with a secondary aim to examine preliminary indications of treatment effect. Accordingly, the determination of the sample size is not based on a statistical power calculation. A sample size of 30 participants is commonly recommended in pilot research as it balances the need to gather sufficient data for feasibility assessment and preliminary efficacy signals while minimizing resource expenditure [44,45]. This size is considered adequate to identify potential issues in study procedures and estimate parameters to inform the design of a larger, definitive trial [45].

### Setting

The study will be carried out by VIRTU Research Group at the Mental Health Center Copenhagen, Denmark, in collaboration with Center for Neuropsychiatric Schizophrenia Research, CNSR, Mental Health Center Glostrup, Denmark. Assessments on clinical metrics at baseline and post treatment will take place at VIRTU Research Group, while EEG examinations will be carried out at CNSR. Both interventions (control and experimental) will be conducted by trained therapists experienced in psychotherapy for psychosis.

### Procedure

Clinical staff from outpatient facilities in the Capital Region of Denmark and Region Zealand will inform eligible participants about the study. Interested participants will be referred to the study by their clinicians. Additionally, participants can contact staff from the study directly to express interest.

After inclusion in the study, the participants will undergo a baseline assessment that involves both clinical interviews, completion of questionnaires, and EEG examinations. Following baseline assessments, participants are randomized to one of the two intervention arms. The assessment procedure is repeated after the intervention; approximately 12 weeks following inclusion. All assessors will receive adequate training prior to conducting clinical assessments and EEG examinations.

Participants in the experimental group (N=15) will be invited to participate in an individual qualitative interview upon treatment completion. The interviews will be conducted by an interviewer not involved in the outcome evaluations. In addition, a group interview with the therapists conducting the experimental intervention will be carried out.

### Interventions

Both interventions will use a manualized format comprising eight individual sessions based on the manual used in the Challenge trial [23]. Drawing on experiences from the Challenge trial, some modifications have been made to the original manual to enhance its flexibility and relevance to the current study. Specifically, greater flexibility has been introduced in transitioning between the different phases of the therapy, allowing sessions to be more responsive to participants’ needs.

To ensure the control group provides an appropriate comparison for the experimental intervention, additional adjustments have been implemented. First, the manual has been extended from seven to eight sessions for both groups. This extension accommodates the additional time required in the experimental group for introducing and utilizing biofeedback. Biofeedback is designed to enhance participants’ awareness of their emotional responses and improve their ability to regulate the distress associated with AVH. Consequently, an explicit focus on emotion regulation has also been incorporated into the therapy manual in the control group. These modifications have been implemented in both groups to ensure that any potential therapeutic effects of the enhanced focus on emotional regulation are accounted for.

### Control group: VR-assisted therapy

The first session focuses on psychoeducation about AVH, including the emotional reactions often associated with hearing voices and commonly used emotion regulation strategies for managing distressing voices. Participants will receive printed materials covering these topics, which they and their relatives can reference throughout the course of the therapy. In the second therapy session, the participant will create an avatar of the hallucinated voice that they decide to work on in therapy, typically corresponding to the most dominant voice (Fig 2). Using specialized computer software, an avatar is created to visually correspond to the participant’s perception of the personification of the voice. Additionally, the voice of the therapist is transformed in a voice transformation program to match the participant’s experience of the speech of the hallucinated voice. In the following sessions, the participant will wear a VR-headset and engage in a virtual dialogue with the avatar. The therapist supports the interaction by alternating between speaking through the voice of the avatar and the voice of a supportive therapist. In the beginning of therapy, the participant is exposed to negative verbatim content from the avatar based on typical content from the hallucinated voice. During this exposure, the therapist supports the participant in confronting the avatar in more assertive manner. Throughout therapy sessions, the avatar will gradually show more compassion towards the participant. The therapeutic aspirations involve enhancing the patient’s sense of control and power over the voice, to strengthen self-esteem, and foster the experience of having a more balanced and equal relationship with the voice.

**Fig 2.**
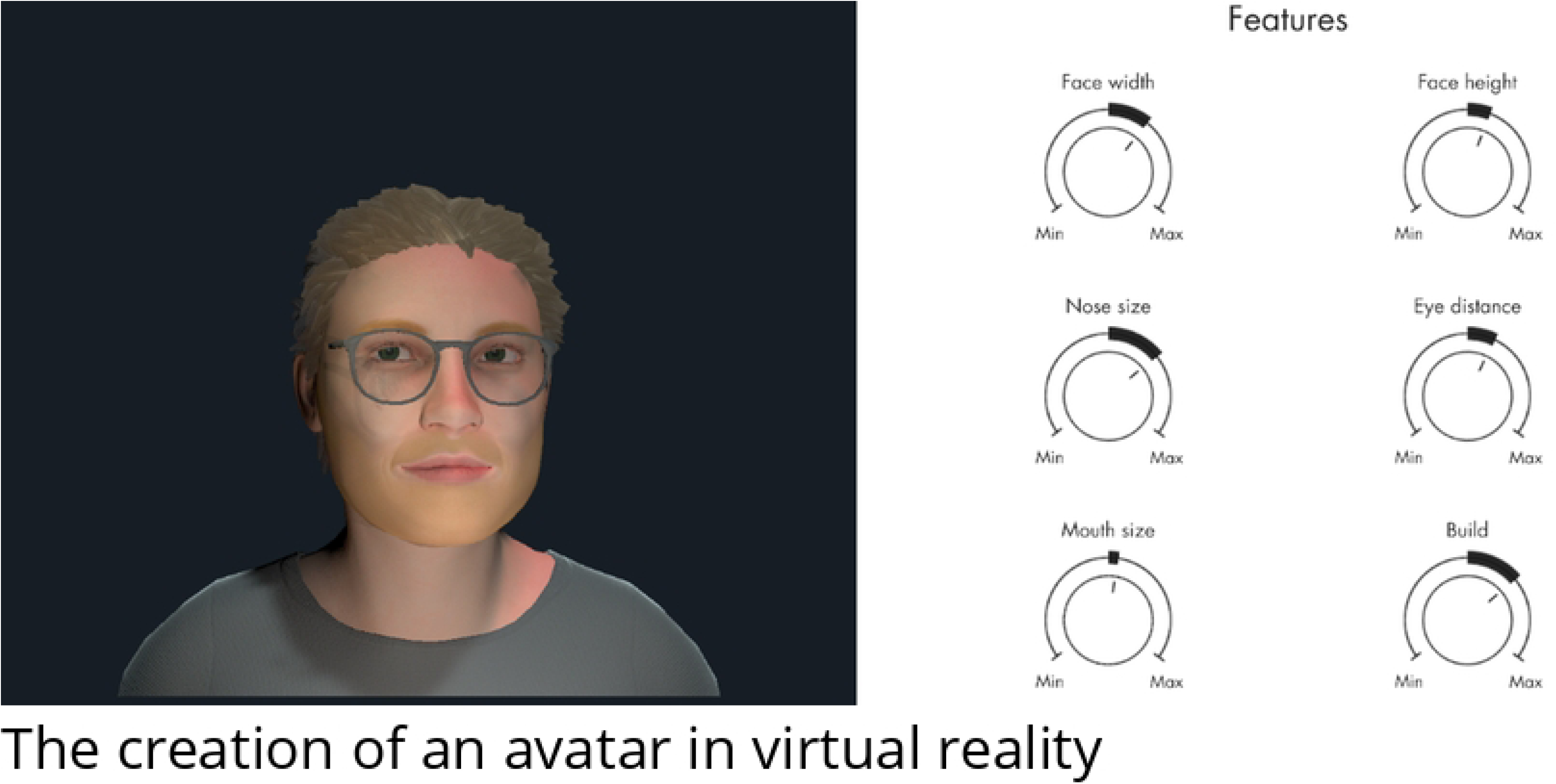
The creation of an avatar in Virtual Reality. An avatar is created to correspond to the participant’s experience of the hallucinated voice by adjusting multiple features in a VR software program, developed by HekaVR.

Each therapy session will start with a therapist-guided exercise focusing on grounding techniques to support the participant in regulating potential anxiety-related reactions prior to the exposure to the avatar. Additionally, existing anxiety regulation techniques are discussed prior to the exposure, enabling the therapist to guide the participant in applying these strategies during the therapy.

### Experimental group: VR-assisted therapy augmented with real-time biofeedback

The experimental group will follow the same therapeutic approach and use the VR-software described in the “control group” but augmented with real-time biofeedback. Biofeedback focuses on heart rate (HR) and heart rate variability (HRV), indicators of autonomic nervous system activity, to provide both participants and therapists with objective, moment-to-moment information about anxiety-related responses.

Biofeedback is delivered via sensors attached to two fingers on one hand (Fig 3).

**Fig 3.**
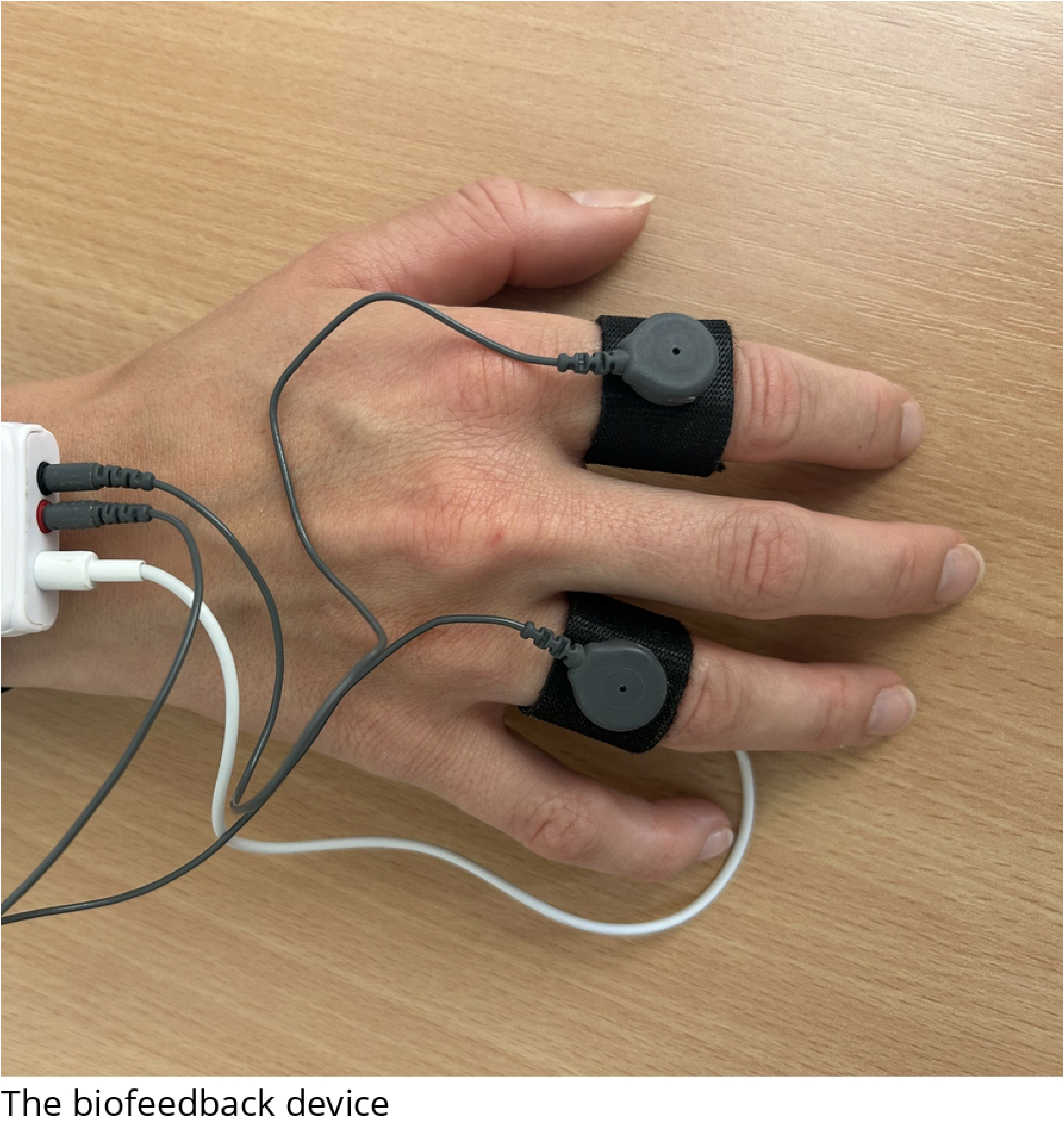
The biofeedback device. The participant will wear Shimmer GSR+ while exposed to the avatar in VR.

In the first therapy session, participants are introduced to the equipment, encouraged to share any concerns, and familiarized with its purpose to ensure comfort before avatar-based work begins. In all subsequent sessions, participants wear the sensors while a baseline HR and HRV recording is obtained during a guided grounding exercise in a calming VR beach environment (15 seconds).This procedure aims to ensure that the participant is as relaxed as possible during the baseline recording. The recording will last for 15 seconds. Following the baseline recording, while the participant is confronted with the avatar, they will receive biofeedback representing changes in the targeted activity compared to the activity recorded at baseline.

During avatar confrontation, changes in HR and HRV relative to baseline are displayed as a thermometer-like visual cue in the VR environment. A blue bar rises or falls from a central line to indicate increases or decreases in physiological arousal. This feedback is also shown in the second session, during avatar creation on a desktop screen, to help participants refine the avatar’s realism while becoming comfortable with the device.

The real-time biofeedback serves two overall purposes in the therapy: It will provide the therapist with an additional and potentially more objective source of information about the participant’s mental state, which will be used to continuously adjust the level of exposure to the avatar. The level of exposure can be adjusted by varying the verbatim content from the avatar and the participant’s distance to the avatar in VR. Second, throughout the therapy session, the participant will be instructed to remain attentive to the feedback display. This will be used as a tool to support the participant in identifying emotional reactions related to the avatar and to enhance awareness of their ability to regulate the emotional distress, supporting an understanding of being more in control.

### Outcomes

#### Primary outcome: Feasibility and acceptability

Primary outcomes include feasibility and acceptability of the experimental intervention and are defined as:

1. A recruitment rate of ≥ 80 % of the target sample in 9 months
2. A retention rate of ≥ 80 % to the study protocol of the recruited participants in the experimental group at cessation of therapy (eight sessions)
3. ≥ 80 % of the participants in the experimental group reporting a satisfaction rating of ≥ 7 on a Likert scale ranging from 1-10 with higher scores indicating a higher level of satisfaction with treatment

#### Secondary outcomes: Indications on treatment effect

For secondary outcomes, the following measures are assessed at baseline and post treatment at 12 weeks follow-up:

- Level of AVH is measured with the Psychotic Symptom Rating Scales, Auditory Hallucinations subscale (PSYRATS-AH) [46]. Outcome measures will include the PSYRATS-AH total score, as well as the two subscales: PSYRATS-AH-Frequency and PSYRATS-AH-Distress.
- Level of acceptance of an action in relation to hallucinated voices is measured with the self-report Voices Acceptance and Action Scale, specifically the VAAS-Action subscale [47]
- Level of assertive responding to voices is measured with the self-report Assertive Responding to Voices – the assertive subscale (Approve – Voices questionnaire) [48].
- Level of perceived power in relation to hallucinated voices is measured with the self-report Voice Power Differential Scale (VPDS) [49]. The total score will be reported as the main measure, and additionally, assertiveness and power items are separately reported.
- Beliefs about voices are measured with the self-report Revised Beliefs about Voices Questionnaires (BAVQ-R) including the Malevolence-, Benevolence- and Omnipotence subscales [50,51].

#### Exploratory outcomes

Exploratory outcomes include:

- Level of positive symptoms including AVH will be measured by the interviewer-rated Scale for the Assessment of Positive Symptoms (SAPS) [52]
- The interviewer-administered Brief Negative Symptoms Scale (BNSS) will measure level of negative symptoms [53].
- Self-reported level of negative symptoms is measured with the Self-evaluation of Negative Symptoms (SNS) [54].
- Ability to use emotion regulation strategies is measured with the self-report Emotion Regulation Questionnaire (ERQ) [55].
- Level of difficulties in emotion regulation is measured with the self-report Difficulties in Emotion Regulation Scale, short version (DERS-16) [56,57].
- The self-report Brief Core Schema Scales: Beliefs about self and others (BCSS) will measure core beliefs about self and others [58].
- Level of depressive symptoms is measured with the interviewer-administered Calgary Depression Scale for Schizophrenia (CDSS) [59].
- Level of suicidal thoughts is measured with the self-report Suicidal Ideation Attributes Scale (SIDAS) [60].
- Level of perceived childhood trauma experiences is measured with the retrospective self-report Childhood Trauma Questionnaire (CTQ) [61,62].
- Level of perceived self-efficacy is measured with the self-report General Self-Efficacy Scale (GSE) [63].
- Level of quality of life is measured with the self-report WHO well-being index (WHO-5) [64].
- Level of sleep quality and disturbances is measured with the self-report Pittsburgh Sleep Quality Index (PSQI) [65].
- Level of readiness for engaging in therapy is measured with the Readiness for Therapy Questionnaire (RTQ) [66].
- Level of insight into mental illness is measured with the self-report Birchwood Insight Scale (BIS) [67].
- Level of satisfaction with therapy is measured with the self-report Client Satisfaction Questionnaire (CSQ) [68].
- Level of symptoms of cybersickness is measured with the Simulator Sickness Questionnaire (SSQ) [69].
- The interviewer-administered Personal and Social Performance Scale (PSP) will measure daily life functioning [70].
- Level of social functioning is measured with the self-report Social Functioning Scale (SFS) [71].

### Electroencephalogram examinations at baseline and post treatment

EEG recordings will be conducted at baseline and post-treatment to investigate neurophysiological changes associated with VR-assisted therapy. Participants will complete five paradigms commonly used to assess auditory processing deficits in schizophrenia:

- **P50 sensory gating:** Measures suppression of the P50 response to paired auditory clicks, reflecting sensory gating [72–74].
- **Mismatch Negativity (MMN):** Captures automatic detection of auditory deviations in frequency or duration, presented while participants watch a muted video to maintain attention[75,76].
- **Selective Attention (SA):** Assesses attentional control using a deviant-tone detection task, alternating between ears [77].
- **40 Hz Auditory Steady-State Response (ASSR):** Evaluates gamma-band oscillatory responses to 40 Hz click trains [78,79].
- **Resting State:** Measures spontaneous EEG activity with eyes open and gaze fixed [80]. These paradigms were selected based on previous studies showing consistent impairments in early auditory processing in schizophrenia that are thought to contribute to delusional beliefs, hallucinations and cognitive and negative symptoms [81–85].

These paradigms were chosen for their robust associations with schizophrenia-related impairments in sensory gating, auditory processing, attention, and oscillatory activity [81–85]. Existing literature supports a link between the event-related potentials (ERPs) P50 and MMN and cognitive dysfunction [82,84]. MMN and ASSR are established biomarkers linked to cognitive dysfunction and treatment response prediction [81,82,84,86–90]. Similar to the MMN, the 40-Hz ASSR, is robustly found to be impaired in patients with schizophrenia [81,89]. Resting-state analyses further capture neural oscillatory abnormalities relevant to psychosis disorder [91,92]. The primary analysis will focus on P50.

Recordings will use a 64-channel BioSemi ActiveTwo System (Amsterdam, Netherlands) with electrodes arranged per the extended 10–20 system and a 2048 Hz sampling rate. Participants will be seated in a sound-insulated cabin (40 dB), and auditory stimuli will be delivered binaurally via insert earphones (E-ARTONE 3A) using Presentation® software (Neurobehavioral Systems, Berkeley, CA). Preprocessing and analysis will follow standard pipelines, adapted for each paradigm.

### Qualitative data

Semi-structured individual interviews with participants in the experimental group will be conducted following completion of treatment, as well as a group interview with the therapists delivering the intervention. The interviews will involve questions regarding:

- Experiences with the biofeedback equipment, which involve the participants’ experience of wearing the watch during therapy sessions, as well as their experience with the technical aspects.
- Experiences with the visibility of the feedback display in the VR environment.
- Experiences with receiving a direct measure of anxiety-related responses during therapy, which involve topics such as the role of biofeedback in supporting the therapeutic aims and sharing these neurophysiological responses with the therapist.

### Randomization and blinding

A computerized randomization system in the Research Electronic Data Capture (REDCap) [93] is used to randomly allocate participants to the two interventions. To equalize participants in both arms, an external party has created and uploaded a block randomization list in REDCap. The block sizes are unknown to the assessors conducting the outcome evaluation and the therapists. The randomization is unstratified. Study therapists, who are not blinded to the intervention allocation, will be responsible for the randomization process. Assessors are blinded to intervention allocation until statistical analyses are completed.

Participants will be instructed to conceal their treatment allocation from the assessors both at baseline assessment, and follow-up assessment.

## Apparatus

### VR headset and software

To enhance the participant’s experience of being immersed in the VR-environment, we will use an Oculus Quest 2 VR headset and noise-cancelling headphones. Audio-visual VR presentation software programs for both intervention groups were developed by HekaVR A/S (Copenhagen, Denmark).

### PPG device used for anxiety monitoring

The selection of the device for measuring physiological signals was based on criteria such as accuracy, reliability, ease of use and patient comfort. HR and HRV data are collected during therapy sessions using the Shimmer3 GSR+ Unit. This unit utilizes the non-invasive methos photoplethysmography (PPG), an optical technique that estimates heart rate based on blood volume changes in the microvascular tissue [38], to obtain these measures. This is a biosensing technique is compatible with the VR setup and have previously been used to assess anxiety levels [38,94,95].

The device is programmed to calculate a Stress Index in real time, combining elevated HR and reduced HRV to reflect sympathetic dominance. HR is normalized to a 0–1 range based on a reference interval of 40–120 bpm, while HRV (lnRMSSD) is inverted and normalized so that lower HRV increases the score. The two components are then weighted and combined into a single value. Higher scores indicate elevated physiological arousal, and the metric is continuously updated and displayed to both the therapist and the participant within the VR software.

Importantly, many on-body biosensors work best when the participant stays stationary [96], which is overall the case for participants in this study. However, noise introduced by potential movement and electrical interferences of the head-mounted display will be addressed by strategies such as filtering and normalization, as well as with a multi-modal approach [38] including 2 different physiological measures.

## Analyses

### Statistical analysis

Feasibility of trial procedures will be assessed by calculating proportions and 95% exact Clopper Pearson confidence intervals covering aspects such as recruitment, consent, participation, randomization, satisfaction with therapy and treatment retention. Indications of treatment effect will be analyzed as a comparison of outcome measures between the two groups using a repeated measures analysis of covariance (ANCOVA), adjusting for baseline differences.

### Analysis of EEG data

The EEG data will be down-sampled and filtered for artifact removal. The exact parameters depend on the specific paradigm. Outcome measures before and after the intervention will be analyzed with repeated-measures analysis of variance. From the P50 paradigm, the response in temporal domains (S1 and S2 amplitude, and the P50 ratio: S2/S1) will be extracted [97]. From the MMN paradigm, the MMN and P3a component from the three different deviants will be extracted. Likewise, the N100, P200, P300, MMN, and PN (processing negativity) will be extracted from the SA paradigm. The 40 Hz ASSR will be analyzed using time-frequency decomposition and the power and inter trials phase coherence around 40Hz will be extracted. Finally, the spectral power of five canonical frequency bands (delta, theta, alpha, beta, and gamma) will be extracted from the resting-state paradigm.

### Qualitative analysis

Data gathered from the individual semi-structured interviews with participants from the experimental group and the group interview with the therapists will be organized and interpreted using a thematic analysis [98]. The electronic qualitative data management system NVivo will be used for the analysis.

### Data management

The study is registered at the Danish Data Protection Agency (p-2024-15496) and at ClinicalTrials.gov (NCT06628323). The study, including all technical solutions, will comply with the General Data Protection Regulation (GDPR). Assessors conducting interviews with the participants will enter the data directly into an electronic case report form (CRF) using REDCap. When necessary, the data will be collected on paper and later entered in REDCap. Each participant’s data is connected to a unique serial number, and only assigned researchers have access to the data in REDCap. Questionnaires can be directly sent from REDCap to the secured public Danish mailing system (E-Boks) that is connected to each participant’s personal identification number (CPR), and completed forms are returned digitally.

Data on paper, consent forms and other physical material with personal information are stored locally and secured. EEG data and audio recordings will be saved on a logged network drive controlled by the Capital Region of Denmark, CIMT, and only assigned researchers and therapists involved in the study have access to the files.

### Ethics

The study is approved by the National Committee on Health Research Ethics for the Capital Region of Denmark (H-24010871). Any important protocol modifications will be submitted to the committee for approval, and relevant updates will be communicated to participants and clinical teams as appropriate. Written informed consent will be obtained from all participants after they have been given oral and written information about the study. The participants are informed that their participation is voluntary and that they can withdraw from the study at any time. Side effects and adverse events will be monitored and documented throughout the study period, and any adverse events assumed to be related to the study will be reported to the Committee on Health Research Ethics of the Capital Region of Denmark. A possible side effect of VR is symptoms of cybersickness, however, VR-assisted therapy is generally considered tolerable for individuals with schizophrenia [99]. Based on this, we do not expect any adverse events.

Positive, negative, and inconclusive results will be disseminated in peer-reviewed scientific journals and will be presented at relevant national and international conferences.

## Discussion

The proposed pilot study aims to evaluate the feasibility, acceptability, and potential treatment efficacy of an innovative treatment approach that combines VR technology with biofeedback to address AVH in individuals with schizophrenia. While VR-based therapy has shown promising potential in reducing the severity of AVH [100,101], to date only one ongoing study investigates the combination of VR with feedback mechanisms in the treatment of AVH, specifically focusing on neurofeedback [41]. The integration of VR with real-time biofeedback, as applied in the present study, remains largely unexplored. Biofeedback augmentation may additionally enhance treatment personalization by enabling real-time continuous adjustments of the intervention, based on individual physiological responses associated with the emotional distress elicited by the hallucinated voice. Moreover, by directly showing participants that they can potentially regulate their emotional responses during the intervention, the biofeedback supplement may enhance their sense of control and empowerment over the voice, which is an important goal of the therapy [15,18,25].

This novel, combined treatment approach builds on a previously studied VR-assisted intervention shown to benefit individuals with AVH [25]. The treatment manual for the current study is developed by therapists experienced with the original intervention, ensuring that any modifications made to integrate the biofeedback component directly address potential challenges faced by this population. Together, these refinements aim to enhance the motivation of participants and referring clinicians and reduce the likelihood of treatment discontinuation.

Despite demonstrated clinical benefits of VR-assisted therapy for auditory verbal hallucinations (AVH), the precise therapeutic mechanisms behind VR-assisted therapy remain unclear. Understanding these mechanisms is crucial for optimizing treatment protocols and tailoring interventions to individual patients [15,102]. Emerging evidence suggests that affect regulation may play a central role by helping patients manage the emotional distress associated with AVH [15,18,19]. However, systematic investigation of affect regulation as a core therapeutic mechanism within VR-based interventions for AVH is still lacking. The present study aims to contribute with new knowledge on whether and how affect regulation functions as a key underlying therapy process in VR-assisted treatments for AVH.

A further strength of the study is the integration of multiple data sources to evaluate both the treatment and study protocol. Pre- and post-treatment EEG examinations will provide valuable insights into neural mechanisms underlying AVH, which will enhance the understanding of the subgroups of participants who respond differently to the treatment.

The findings from this pilot study will provide crucial insights into the use of physiological markers in guiding VR-assisted interventions for AVH and determine whether this approach warrants further exploration. If successful, it could lay the groundwork for larger clinical trials aimed at documenting the effect of this augmentation.

### Status of the study

The recruitment of participants started in November 2024 and is ongoing.

## Authors contribution

AFH and SBS share first authorship of the manuscript. AFH has written the first draft of the manuscript under supervision of LBG. SBS has provided critical contribution in reviewing and editing the final manuscript. ASD and THT are study therapists that have contributed to the development of the treatment protocols for the study.

KML has contributed as a counselor in EEG data. BHE, TDK and KSA have contributed to the design of EEG data collection. JDC has contributed to device evaluation and EEG procedure documentation. KE, DL and KWM have contributed to conceptualizing the integration of biofeedback. MN has contributed to the design of the study. LBG is the initiator and principal investigator of the study and is the primary contributor to the conceptualization and methodology of the study. All authors revised and approved the final manuscript.

## Data Availability

No datasets were generated or analysed during the current study. All relevant data from this study will be made available upon study completion.

## Acknowledgements

The authors express their immense gratitude to all the participants of the study and the psychiatric clinics that will support with recruiting participants. We would also like to thank HekaVR for their work on integrating the biofeedback into the VR software.

## Author note

The avatars depicted on the figure 2 are created by the study staff.

## Supporting information

**S1 File. SPIRIT checklist.** Completed SPIRIT 2025 checklist indicating where each item is addressed in the manuscript.

**S2 File. Latest protocol approved by ethics committee.** The latest version of the protocol approved by the National Committee on Health Research Ethics for the Capital Region of Denmark (Version 5.0, 14. April 2025, H-24010871).

**S1 Fig. CONSORT 2025 Flow diagram.** Schedule showing the timing of participant enrolment, interventions, and assessments.

